# VIsual STAndardized Quantification of LGE (VISTAQ), a contour-less method for late gadolinium enhancement quantification

**DOI:** 10.64898/2026.04.09.26350552

**Authors:** Giovanni Donato Aquaro, Roberto Licordari, Carmelo De Gori, Giancarlo Todiere, Umberto Ianni, Andrea Barison, Antonio De Luca, Alessandro Folgheraiter, Crysanthos Grigoratos, Mattia Alberti, Lisa Fulceri, Marilena Lombardo, Raffaele De Caterina, Gianfranco Sinagra, Michele Emdin, Gianluca Di Bella

## Abstract

**Background:** Late gadolinium enhancement (LGE) quantification by cardiovascular magnetic resonance is central to risk stratification in hypertrophic cardiomyopathy (HCM), yet conventional techniques require contour tracing and region-of-interest (ROI) placement, which may reduce reproducibility and increase analysis time. We developed a novel visual standardized approach, the Visual Standardized Quantification of LGE (VISTAQ), that does not require myocardial contouring, arbitrary ROI positioning, or dedicated post-processing software.

**Methods:** In this multicenter, multivendor retrospective study, LGE images from 400 patients (100 prior myocardial infarction, 250 HCM, 50 other non-ischemic heart diseases) were analyzed. VISTAQ subdivides each myocardial segment into transmural mini-segments and classifies LGE visually using predefined criteria, expressing global LGE burden as the percentage of positive mini-segments. Reproducibility was assessed in 250 patients across different observer expertise levels using intraclass correlation coefficients (ICC) and Bland–Altman analysis. In 100 HCM patients, VISTAQ was compared with conventional methods (mean+2SD, +5SD, +6SD, FWHM, visual thresholding). Prognostic performance was evaluated in 250 HCM patients over a median 5-year follow-up.

**Results:** VISTAQ demonstrated excellent intra- and inter-observer reproducibility (ICC up to 0.98 and 0.97, respectively), consistent across disease subtypes. Compared with conventional techniques, VISTAQ showed similar ICC to FWHM but significantly lower net and absolute inter-observer differences (median absolute difference 1.3%). Mean+2SD markedly overestimated LGE, whereas mean+6SD slightly underestimated LGE compared with VISTAQ, mean+5SD, FWHM, and visual thresholding. Analysis time was substantially shorter with VISTAQ (median 105 vs. 375 seconds, p<0.0001). During follow-up, 21 hard cardiac events occurred in HCM population. An LGE threshold >10% predicted events with higher accuracy using VISTAQ (AUC 0.90; sensitivity 85%; specificity 94%) compared with mean+6SD (AUC 0.75; sensitivity 57%; specificity 93%).

**Conclusions:** VISTAQ provides highly reproducible, time-efficient LGE quantification without dedicated software and demonstrates non-inferior prognostic discrimination in HCM compared with conventional threshold-based techniques.

## Introduction

Late gadolinium enhancement (LGE) imaging is one of the cornerstone techniques in cardiovascular magnetic resonance (CMR). It enables robust tissue characterization and allows differentiation between ischemic myocardial injury, typically following coronary artery disease, and non-ischemic patterns of damage as seen in myocarditis and various cardiomyopathies (1). Beyond its diagnostic value, LGE burden has a well-established prognostic impact across a broad spectrum of cardiac diseases. In hypertrophic cardiomyopathy (HCM) in particular, quantitative assessment of LGE is essential, as current European Society of Cardiology (ESC) guidelines recognize an extent ≥15% of left ventricular mass as a marker associated with adverse outcomes (2).

Despite this central role, the optimal method for LGE quantification remains debated. Current guideline recommendations endorse a signal intensity threshold defined as the mean plus 2 standard deviations (SD) of a region of interest (ROI) placed in visually normal myocardium (2). However, multiple studies have demonstrated that this approach is inaccurate (3–5). In inversion-recovery LGE sequences, normal myocardium is effectively nulled, and its residual signal mainly represents background noise, which does not follow a Gaussian distribution as typically assumed for parameters of human populations (3). Alternative methods, such as a threshold of mean + 6SD, lack a strong physiological rationale but tend to match more closely the visual assessment of experienced readers. The full-width-at-half-maximum (FWHM) technique is another commonly used approach, yet it is limited by the need to place an ROI within areas of enhancement and performs suboptimally in the presence of small, heterogeneous, or patchy scars (4). All these techniques require manual delineation of endocardial and epicardial borders and the placement of ROIs in presumed normal (or enhanced) myocardium, steps that substantially increase intraobserver variability and may undermine the reproducibility of LGE quantification.

In this study, we proposed a novel LGE quantification approach, the Visual Standardized Quantification of LGE (VISTAQ), based on a structured visual assessment that did not require manual contouring of the myocardium nor arbitrary placement of ROIs in either remote or enhanced regions and did not require post-processing software. Specifically, we: (a) compared the performance of VISTAQ against the conventional methods for LGE quantification; (b) assessed the intra- and inter-observer reproducibility of VISTAQ; and (c) evaluated and compared the prognostic value of VISTAQ and the mean + 6SD technique in a cohort of patients with hypertrophic cardiomyopathy.

## Methods

In this multicenter, multivendor study we retrospectively analyzed LGE images from patients with prior myocardial infarction (occurred >6 months before CMR), patients with hypertrophic cardiomyopathy (HCM) and patients with other causes of non ischemic heart disease.

HCM was defined as a maximal left ventricular (LV) wall thickness ≥15 mm in one or more LV myocardial segments in the absence of secondary causes of LV hypertrophy, with the exception of patients with systemic hypertension under effective pharmacological treatment who also had a family history of HCM or a consistent pathogenic genotype (2). Patients with suboptimal LGE image quality were excluded.

### CMR acquisition protocol

CMR examinations were performed on 1.5-T scanners from different vendors (Signa HDx, GE Healthcare, Milwaukee, WI, USA; Magnetom Avanto, Siemens Medical Systems, Erlangen, Germany; Gyroscan NT, Philips Healthcare, Amsterdam, The Netherlands) using dedicated cardiac coils. The study protocol included functional assessment with conventional short-axis cine images acquired from the mitral valve plane to the LV apex using a steady-state free precession pulse sequence. Additional cine images with identical parameters were acquired in standard 2-, 3-, and 4-chamber views.

LGE images were acquired 10 minutes after administration of a gadolinium-based contrast agent at a dose of 0.2 mmol/kg in contiguous short-axis slices. An inversion-recovery T1-weighted fast gradient-echo sequence was used with the following typical parameters: field of view 350–400 mm, slice thickness 8 mm, 0-mm interslice gap, repetition time 3–5 ms, echo time 1–3 ms, flip angle 18–25°, acquisition matrix 224 × 224, and reconstruction matrix 256 × 256. The inversion time was individually adjusted to null normal myocardium using a TI-scout sequence. The use of a phase-sensitive inversion-recovery (PSIR) technique was left to the discretion of the operators at each participating center.

### Quantification of LGE extent by the conventional methods

In each center, late gadolinium enhancement (LGE) images were analyzed by experienced readers with level III cardiovascular magnetic resonance (CMR) accreditation from the European Association of Cardiovascular Imaging (EACVI), who were blinded to all clinical data. LGE extent was quantified using a commercially available research software package (CVI42, Circle Imaging).

Endocardial and epicardial contours were manually traced on each short-axis slice to delineate the left ventricular (LV) myocardium. LGE extent was quantified using five different methods: (1) mean +2 SD; (2) mean +5 SD; (3) mean +6 SD; (4) the full width at half maximum (FWHM) method; and (5) a manual threshold method.

For the mean +2 SD, +5 SD, and +6 SD methods, a region of interest (ROI) was placed in visually normal remote myocardium without detectable LGE to determine the mean signal intensity (SI) and its standard deviation (SD). Myocardial voxels with SI values >2 SD, >5 SD, and >6 SD above the mean of the reference region were classified as enhanced, respectively.

For the FWHM method, an additional ROI was placed in a visually identified region of enhanced myocardium, and the SI threshold for LGE was defined according to previously published methodology.

In the manual threshold method, the SI threshold was manually selected by the investigators by progressively increasing the threshold value using a slider tool and comparing the resulting parametric maps with the original LGE images. The final threshold was defined as the value that most closely matched the visual interpretation of LGE.

The percentage of enhanced voxels within the entire LV myocardium was subsequently calculated. LGE extent was expressed both in grams and as a percentage of total LV mass.

### Visual standardized quantification of late gadolinium enhancement (VISTAQ)

LGE was assessed using the novel method called VIsual STandardized Quantification method (VISTAQ). VISTAQ rules are reassumed in figure 1. Briefly, for each short-axis slice, the left ventricular myocardium was segmented according as follows: 6 segments in each basal slice, 6 segments in each mid-ventricular slice, and 4 segments in each distal slice. In addition, a single “true apex” segment was defined on a long-axis view.

**Figure 1:**
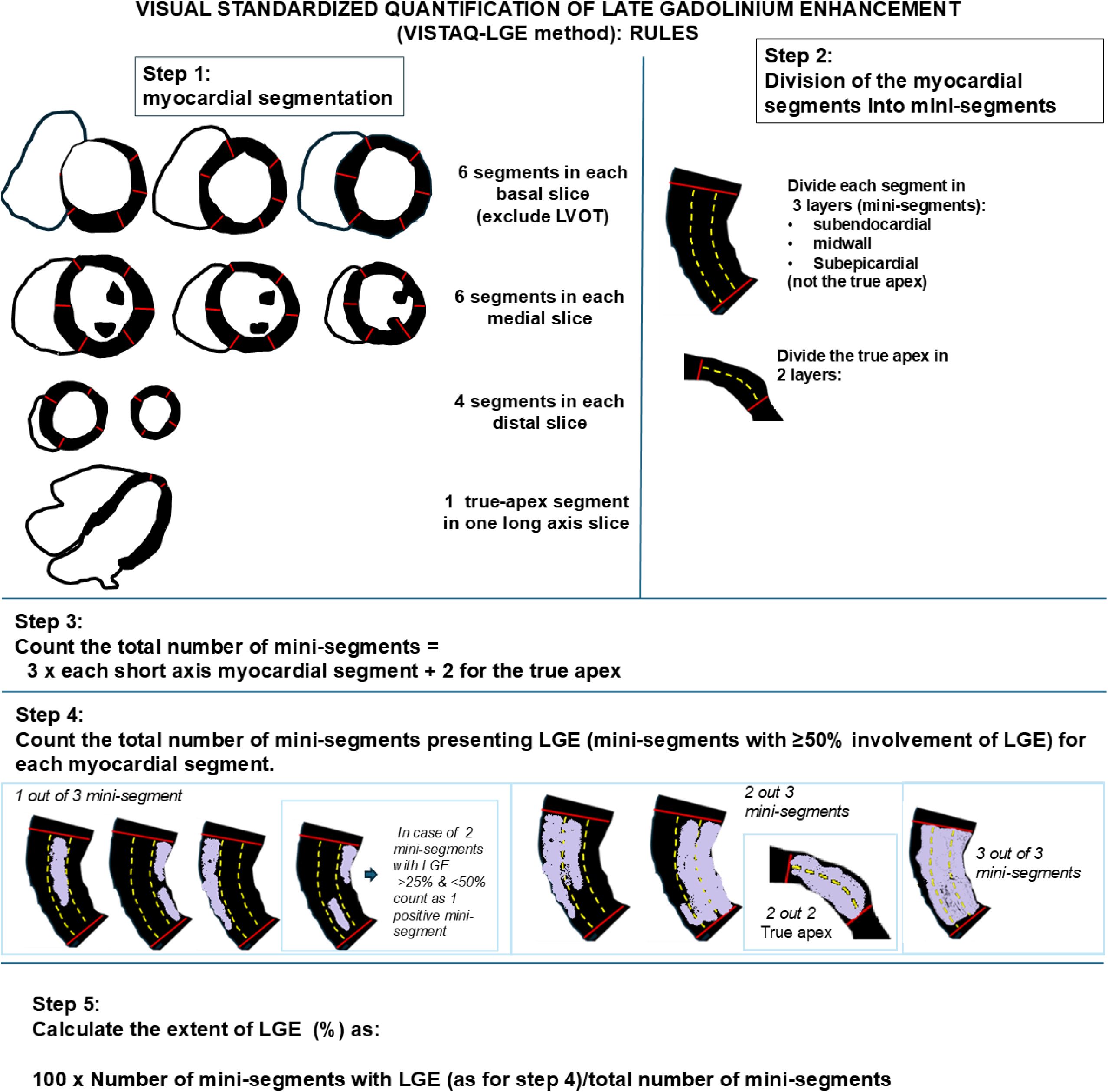
Rules of VISTAQ analysis for quantification of LGE extent. Schematic representation of the VISTAQ method for LGE quantification. Step 1: Myocardial segmentation is performed on all the short-axis slices, dividing basal and mid-ventricular levels into 6 segments each (excluding the left ventricular outflow tract), distal slices into 4 segments, while true apex was considered as 1 segment. Step 2: Each myocardial segment is further subdivided into three mini-segments (endocardial, mid-wall, and subepicardial), while the true apex is divided into mini-segments. Step 3: For each slice, the total number of mini-segments is counted (3 × each short-axis myocardial segment + 2 for the true apex). Step 4: The number of mini-segments showing LGE involvement ≥50% is visually assessed and recorded. Step 5: The extent of LGE is calculated as the percentage of positive mini-segments relative to the total number of evaluable mini-segments.

In a second step, each short-axis myocardial segment was subdivided into three concentric transmural layers (“mini-segments”: subendocardial, mid-wall, and subepicardial), whereas the true apex segment was subdivided into two layers. The total number of mini-segments was therefore calculated as: 3 × (number of short-axis segments) + 2 (true apex). LGE was then visually scored in each mini-segment, which was classified as LGE-positive when ≥50% of its area showed hyperenhancement. When two or more mini-segments within the same segment visually exhibited an LGE extent <50%, but approximately involving one third of each mini-segment, they were collectively counted as one positive mini-segment.

For each myocardial segment, the number of LGE-positive mini-segments (0, 1, 2, or 3 in short-axis segments; 0, 1, or 2 in the true apex) was recorded. The global extent of LGE was finally expressed as a percentage of myocardial involvement, computed as:

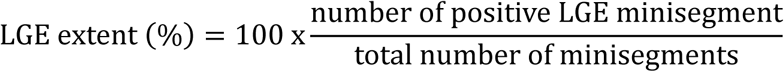

Provided as supplemental material is the file *VISTAQ LGE.html*, which was used to calculate LGE extent using the VISTAQ method. In addition, VISTAQ can be accessed online at https://www.cmr-report.com/VISTAQ/.

### Prognostic evaluation

A structured clinical questionnaire was completed by a physician during scheduled outpatient visits at each participating hospital, by telephone contact with patients’ relatives, or through their general practitioners. The questionnaire recorded the occurrence of hard cardiac events, defined as cardiac death, resuscitated cardiac arrest, appropriate ICD shock, antitachycardia pacing, and sustained ventricular tachycardia documented on Holter ECG monitoring, as well as secondary end points (heart failure hospitalization, stroke, atrial fibrillation, and acute coronary syndrome).

Device interrogations were systematically reviewed by the referring cardiologist to confirm the appropriateness of ICD therapies. The occurrence of hard cardiac events was adjudicated by a panel of expert investigators.

### Statistical analysis

Values are presented as the mean ± SD or as the median (interquartile range) for variables with normal and non-normal distributions, respectively. Normal distribution was assessed using the Kolmogorov-Smirnov test.

Intra- and inter-observer variability for LGE quantification with VISTAQ-LGE was assessed by calculating the intraclass correlation coefficient (ICC). Bland–Altman plots were generated to evaluate intra- and inter-observer agreement across the different groups of readers. Reproducibility was classified as “poor” for ICC values <0.50, “moderate” for values between 0.50 and 0.75, “good” for values between 0.75 and 0.90, and “excellent” for values >0.90.

A receiver operating characteristic (ROC) curve was used to identify the optimal LGE threshold, measured with VISTAQ-LGE method, for predicting cardiac events. Kaplan–Meier time-to-event analysis was performed to estimate and compare event-free survival across groups. Cox regression analysis was used to assess the impact of variables that were significant in univariable analysis on the occurrence of hard cardiac end points. A p-value <0.05 was considered statistically significant.

## Results

### Intra- and inter-observer reproducibility

The evaluation of intra- and inter-observer reproducibility of VISTAQ was performed in a population of 250 patients (100 with previous myocardial infarction, 100 with Hypertrophic cardiomyopathy, and 50 other non-ischemic cardiac diseases). Characteristics of this population are summarized in table 1-3.

**Table 1.**
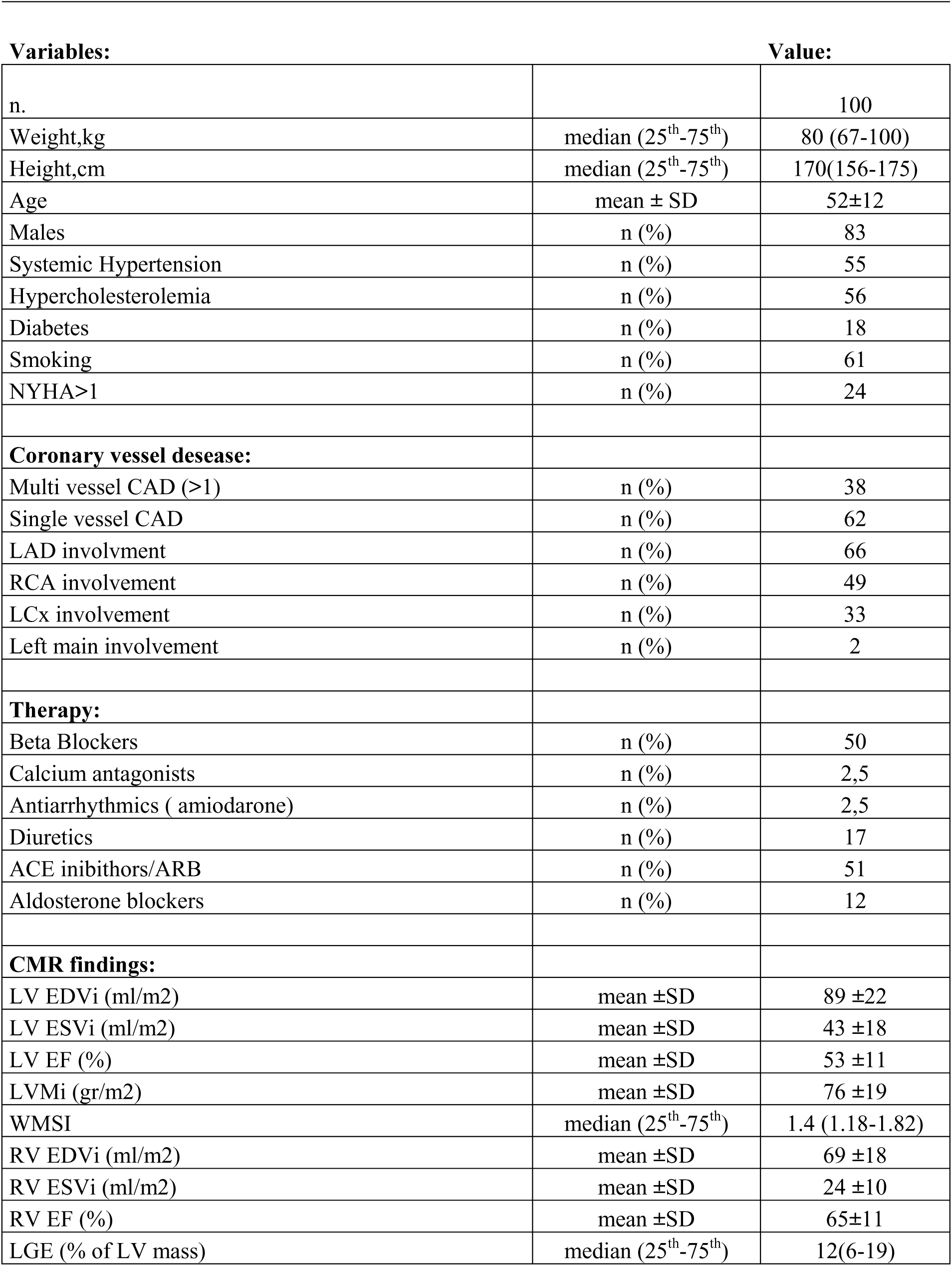
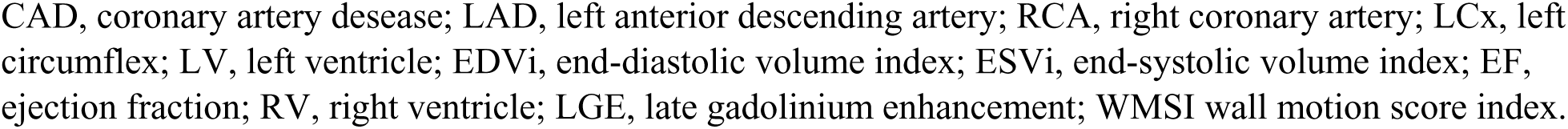
Characteristics of the population with previous myocardial infarction.

**Table 2.** Clinical characteristics of the population of HCM for reproducibility of VISTAQ.

| **Variables:** |  | **Value:** |
| --- | --- | --- |
| n. |  | 100 |
| Age (years) | mean ±SD | 52±15 |
| Males | n (%) | 71 |
| Systemic Hypertension | n (%) | 7 |
| Hypercholesterolemia | n (%) | 16 |
| Diabetes | n (%) | 8 |
| Smoking | n (%) | 10 |
| Dyspnea (NYHA class>I) | n (%) | 35 |
| 5-years HCM risk SCD<4% | n (%) | 85 |
| 5-years HCM risk SCD 4-6% | n (%) | 15 |
| **Therapy:** |  |  |
| Beta Blockers | n (%) | 46 |
| Calcium antagonists | n (%) | 8 |
| Antiarrhythmics | n (%) | 3 |
| Diuretics | n (%) | 12 |
| ACE inibithors | n (%) | 33 |
| Aldosterone blockers | n (%) | 3 |
| **CMR findings:** |  |  |
| Maximal WT≥30 (mm) | n (%) | 4 |
| Family history of SCD | n (%) | 19 |
| Syncope | n (%) | 9 |
| Non-sustained VT | n (%) | 44 |
| Obstructive HCM | n (%) | 20 |
| LV outflow gradient (mmHg) | mean ±SD | 53 ±30 |
| LV atrium (mm) | median (25<sup>th</sup>-75<sup>th</sup>) | 32 (30-43) |
| LV EDVi (ml/m2) | mean ±SD | 69 ± 17 |
| LV ESVi (ml/m2) | mean ±SD | 21 ± 8 |
| LV EF (%) | mean ±SD | 68 ± 12 |
| LVMi (gr/m2) | mean ±SD | 114 ±51 |
| RV EDVi (ml/m2) | mean ±SD | 63 ±17 |
| RV ESVi (ml/m2) | mean ±SD | 20 ±8 |
| RV EF (%) | mean ±SD | 69 ±9 |
| LGE (%) | median (25<sup>th</sup>-75<sup>th</sup>) | 2 (1-9) |

HCM, hypertrophic cardiomyopathy; SCD, sudden cardiac death; WT, wall thickness; VT, ventricular tachycardia; LV, left ventricle; EDVi, end-diastolic volume index; ESVi, end-systolic volume index; EF, ejection fraction, RV, right ventricle; LGE, late gadolinium enhancement.

**Table 3:** Characteristics of the Study Population of Non Ischemic Cardiac disease.

| Characteristic |  |
| --- | --- |
| n. | 50 |
| Males, n(%) | 39(78%) |
| Age, yrs | 42 (22 – 55) |
| Weight, kg | 73 ±15 |
| Height, cm | 171 ±10 |
| **Type of NICD:** |  |
| Previous myocarditis | 30(60%) |
| DCM | 10(20%) |
| NDLVC | 10(20%) |
| **Risk Factors:** |  |
| Hypertension | 1 (2%) |
| Hypercholesterolemia | 1 (2%) |
| Diabetes Mellitus | 1 (2%) |
| Smoking | 2(4%) |
| Family history of CAD | 3 (6%) |
| **CMR data:** |  |
| LVEDVi, ml/m<sup>2</sup> | 89 (68-98) |
| LVESVi, ml/m<sup>2</sup> | 30 (25-38) |
| LV EF, % | 55 (44–66) |
| WMSI >1, n | 10 (21%) |
| LVMi, g/m<sup>2</sup> | 68 (62–70) |
| RVEDVi, ml/m<sup>2</sup> | 80 (66-92) |
| RVESVi, ml/m<sup>2</sup> | 32 (26-39) |
| RV EF, % | 59 (54-69) |
| LGE extent, % of LV mass | 2(0-5) |

In the whole population, quantification of LGE extent by VISTAQ method demonstrated an excellent intra-observer reproducibility (single measures ICC 0.96, 95% CI 0.95-0.97, average measures ICC 0.98, 95% 0.97-0.99), an excellent inter-observer reproducibility (single measures ICC 0.94, 95% CI 0.91-0.95, average measures ICC 0.97, 95% 0.96-0.97).

In the population of patients with previous myocardial infarction, VISTAQ demonstrated an excellent intra-observer reproducibility (single measures ICC 0.94, 95% CI 0.90-0.97, average measures ICC 0.97, 95% 0.95-0.98), and a very good/excellent inter-observer reproducibility (single measures ICC 0., 91 95% CI 0.87-0.93, average measures ICC 0.94, 95% 0.90-0.96). Example of VISTAQ analysis compared to FWHM in a case of previous myocardial infarction is shown in figure 2.

**Figure 2:**
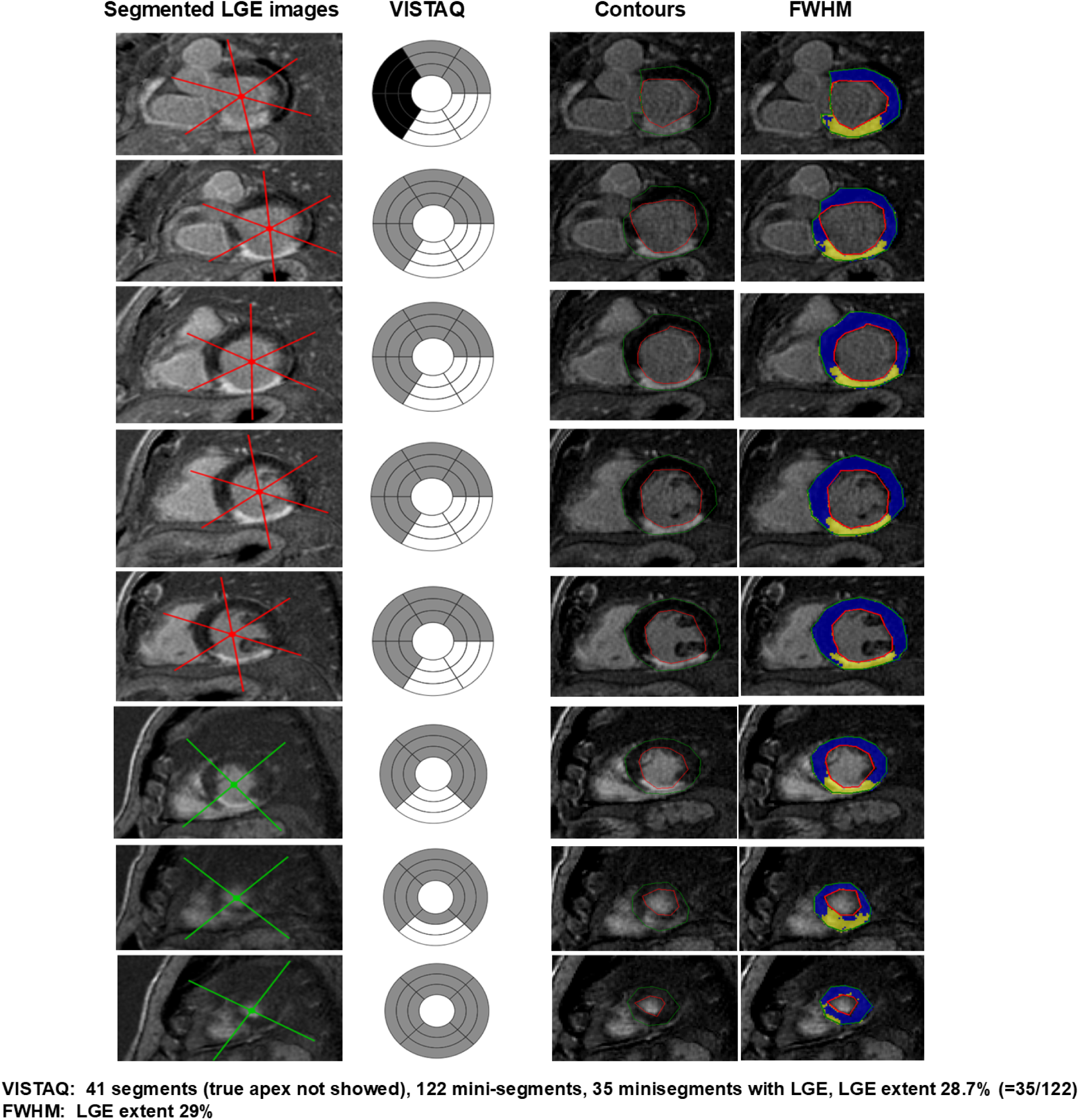
Example of VISTAQ analysis in a patient with previous myocardial infarction. In the first column, original LGE images are shown with myocardial segmentation into 6 segments (basal and mid-ventricular slices) or 4 segments (distal slices). In the second column, VISTAQ analysis was performed on bull’s-eye plots generated from this segmentation. Black indicates mini-segments excluded from analysis because they are located at the level of the LVOT, grey indicates mini-segments without LGE, and white indicates mini-segments with LGE. In this case, a total of 122 mini-segments were analyzed, of which 35 showed LGE, corresponding to an LGE extent of 28.7% of left ventricular mass. In the third column, endocardial and epicardial contours of the same patient’s LGE images are shown, while the fourth column displays the parametric map obtained using the Full Width at Half Maximum (FWHM) method. With this approach, LGE extent was 28.8%.

In the population of patients with HCM, VISTAQ demonstrated an excellent intra-observer reproducibility (single measures ICC 0.92, 95% CI 0.81-0.96, average measures ICC 0.96, 95% 0.89-0.98), and very good/excellent inter-observer reproducibility (single measures ICC 0.91, 95% CI 0.70-0.93, average measures ICC 0.92, 95% 0.82-0.96). Example of VISTAQ analysis compared to mean+6SD in a case of HCM is shown in figure 3.

**Figure 3:**
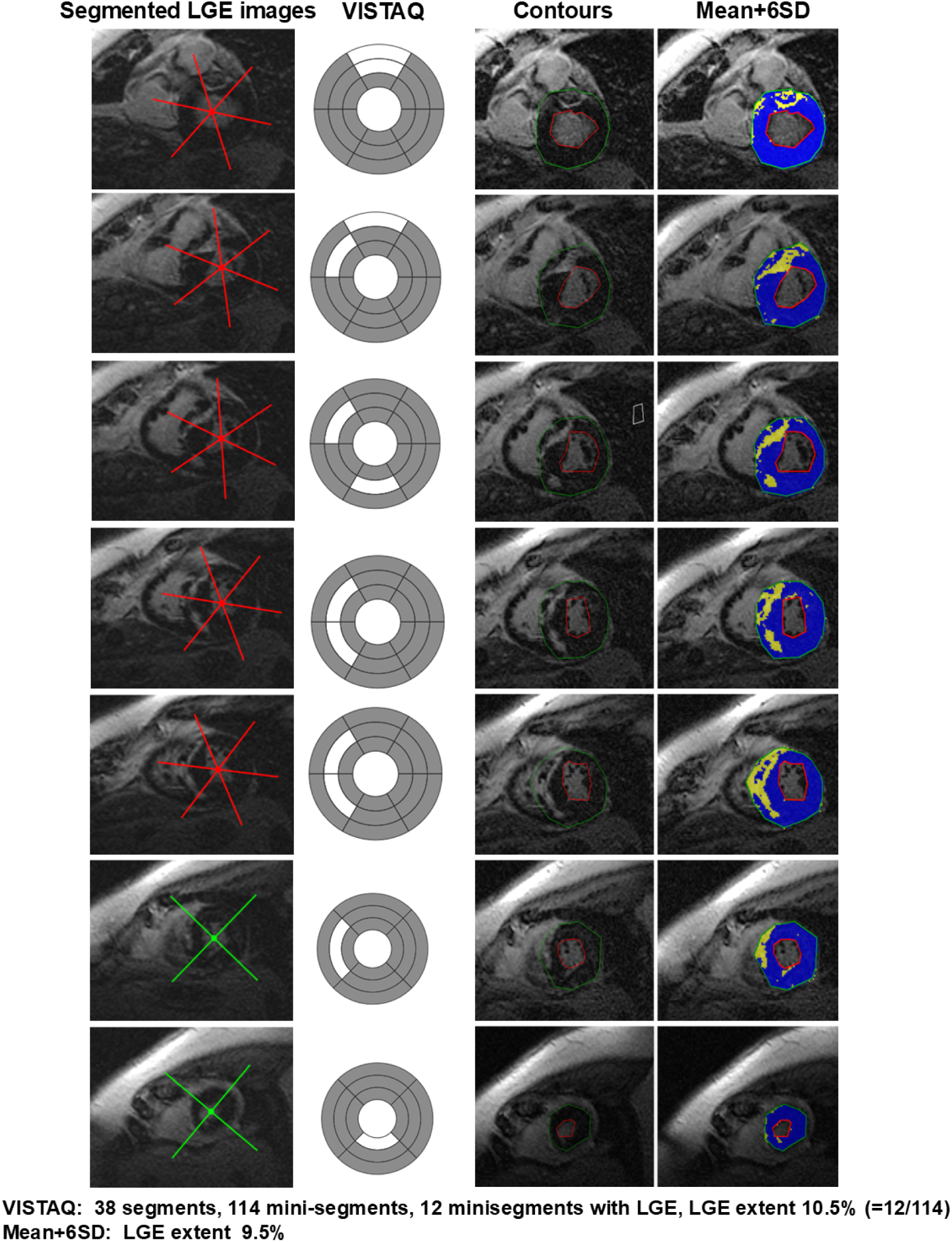
Example of quantification of LGE using mean+6SD and VISTAQ in a patient with HCM.

In the population of patients with non-ischemic LGE without LV-hypertrophy, VISTAQ demonstrated an excellent intra-observer reproducibility (single measures ICC 0.98, 95% CI 0.94-0.99, average measures ICC 0.99, 95% 0.97-0.99), and a very good/excellent inter-observer reproducibility (single measures ICC 0.91, 95% CI 0.81-0.96, average measures ICC 0.95, 95% 0.89-0.98). Example of VISTAQ analysis compared to mean+6SD in a case of previous myocarditis is shown in figure 4.

**Figure 4:**
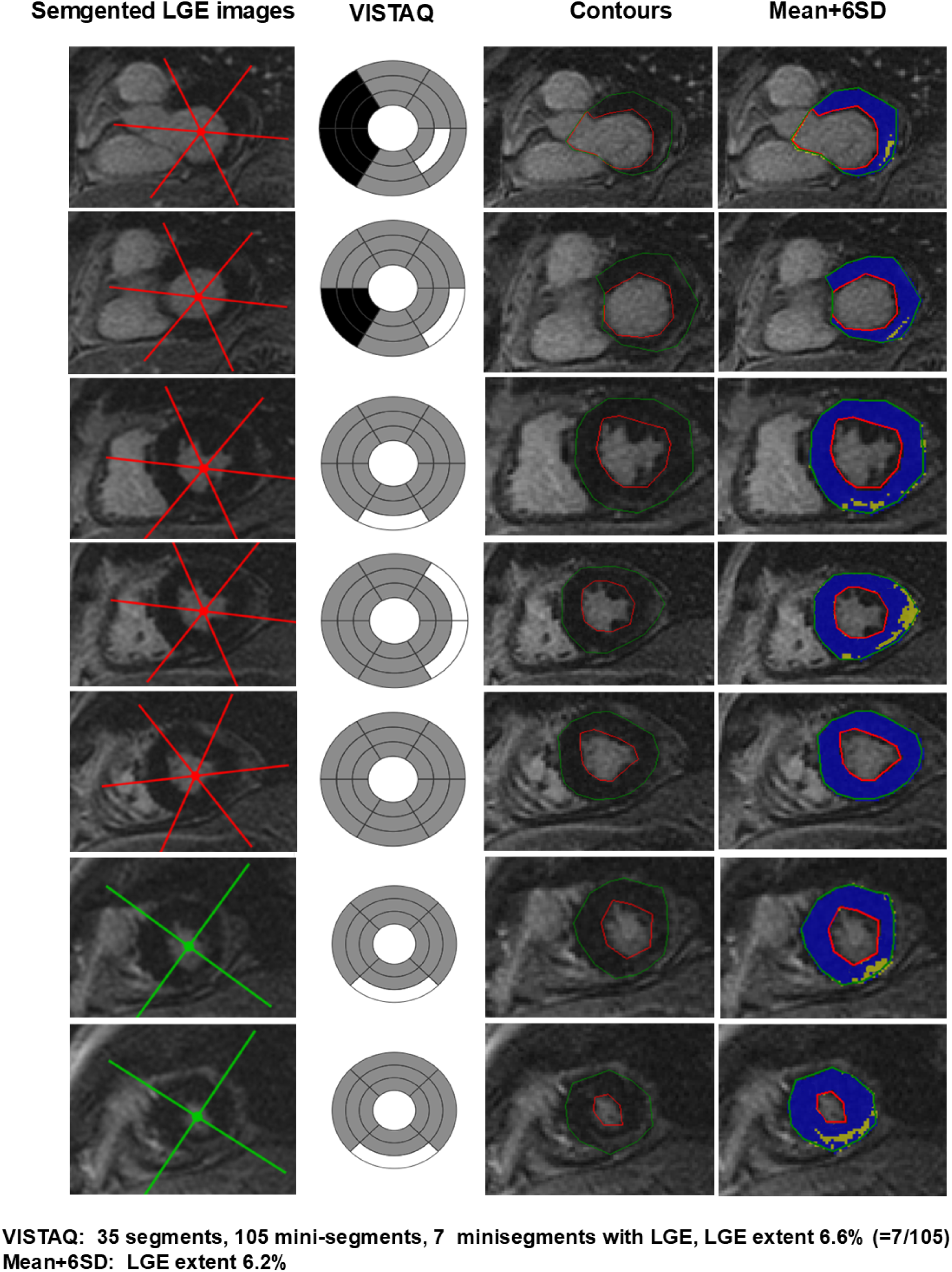
Example of quantification of LGE using mean+6SD and VISTAQ in a patient with non-ischemic fibrosis (previous myocarditis).

### Comparison with conventional methods

The comparison of reproducibility among VISTAQ and other conventional methods for LGE quantification (mean+2SD, +5SD, +6SD, FWHM and manual threshold methods) were performed in 100 patients with HCM. ICC for intra- and inter-observer reproducibility of conventional techniques are shown in table 4.

**Table 4:** Intraclass coefficients.

| **Methods:** | **Group** | **Inter-/Intra-**<br>**observer** | **ICC**<br>**(single)** | **95% CI** | **ICC**<br>**(average)** | **95% CI** |
| --- | --- | --- | --- | --- | --- | --- |
| **VISTAQ** | Whole population | Intra-observer | 0.96 | 0.95-0.97 | 0.98 | 0.97-0.99 |
|  |  | Inter-observer | 0.94 | 0.91-0.95 | 0.97 | 0.96-0.97 |
|  | MI | Intra-observer | 0.94 | 0.90-0.97 | 0.97 | 0.95-0.98 |
|  |  | Inter-observer | 0.91 | 0.82-0.93 | 0.94 | 0.90-0.96 |
|  | HCM | Intra-observer | 0.92 | 0.81-0.96 | 0.96 | 0.89-0.98 |
|  |  | Inter-observer | 0.91 | 0.70-0.93 | 0.95 | 0.82-0.96 |
|  | NI-LGE | Intra-observer | 0.98 | 0.94-0.99 | 0.99 | 0.97-0.99 |
|  |  | Inter-observer | 0.91 | 0.81-0.96 | 0.95 | 0.89-0.98 |
| **Mean+2SD** | HCM | Intra-observer | 0.90 | 0.88-0.95 | 0.93 | 0.90-0.98 |
|  |  | Inter-observer | 0.79 | 0.67-0.95 | 0.94 | 0.80-0.98 |
| **Mean+5SD** | HCM | Intra-observer | 0.90 | 0.87-0.97 | 0.97 | 0.96-0.98 |
|  |  | Inter-observer | 0.78 | 0.75-0.94 | 0.92 | 0.88-0.97 |
| **Mean+6SD** | HCM | Intra-observer | 0.92 | 0.87-0.97 | 0.94 | 0.94-0.97 |
|  |  | Inter-observer | 0.78 | 0.76-0.94 | 0.91 | 0.87-0.97 |
| **FWHM** | HCM | Intra-observer | 0.95 | 0.92-0.96 | 0.97 | 0.96-0.98 |
|  |  | Inter-observer | 0.91 | 0.89-0.98 | 0.94 | 0.89-0.98 |
| **Manual** | HCM | Intra-observer | 0.92 | 0.88-0.94 | 0.94 | 0.89-0.97 |
|  |  | Inter-observer | 0.76 | 0.75-0.97 | 0.90 | 0.85-0.98 |

The median difference of LGE extent between two different investigator was calculated for each method as net difference (including negative numbers) and as absolute difference (used absolute value of negative numbers). VISTAQ method showed both lower net (median of 0, 95% CI -1.8-0.8) and absolute (median 1.3, 95% CI 0.3-3.4) difference than FWHM (median of net difference 2.5, IQR 0.2-5.8,p<0.0001; median of absolute difference 3.1, IQR 1.5-5.8, p<0.0001), mean+2SD (median of net difference 4.8, IQR CI -1.3-9.8, p<0.0001; median of absolute difference 5.2, IQR 2.6-9.6, p<0.0001), mean+5SD (median of net difference -2.2, 95% IQR -4.4-0.4,p=0.008; median of absolute difference 2.8, IQR 1.3-4.7, p=0.007), mean+6SD (median of net difference -1.5, 95% IQR-3.5-0.8, p=0.04; median of absolute difference 2.1, IQR 1-3.8, p=0.02) and visual threshold (median of net difference -2.7, IQR -5.9- -0.6, p<0.0001; median of absolute difference 3.5, IQR 1.8-6.5, p<0.0001).

Overall, The mean+2SD method significantly overestimated LGE extent (median extent 22%, IQR 15-33) compared to mean+5SD (median extent 8%, IQR 4-15, p<0.0001), to mean+6SD (median extent 7%, IR 2-12, p<0.0001), to FWHM (median extent 9%, IQR 5-16, p<0.0001), to visual threshold method (median extent 8%, IQR 6-10, p<0.0001) and to VISTAQ (median extent 9%, IQR 5-15, p<0.0001).

On contrast, LGE extent by mean+6SD method was significantly lower than by mean+5SD (p=0.02), by FWHM (p=0.002), by visual threshold (p=0.002) and by VISTAQ (p=0.002). No significant differences were found when LGE extent was measured by mean+5SD, FWHM, visual threshold and VISTAQ.

Bland-Altman plots for intra- and inter-observer agreement for all the methods of LGE quantification are shown respectively in figure 5 and 6.

**Figure 5:**
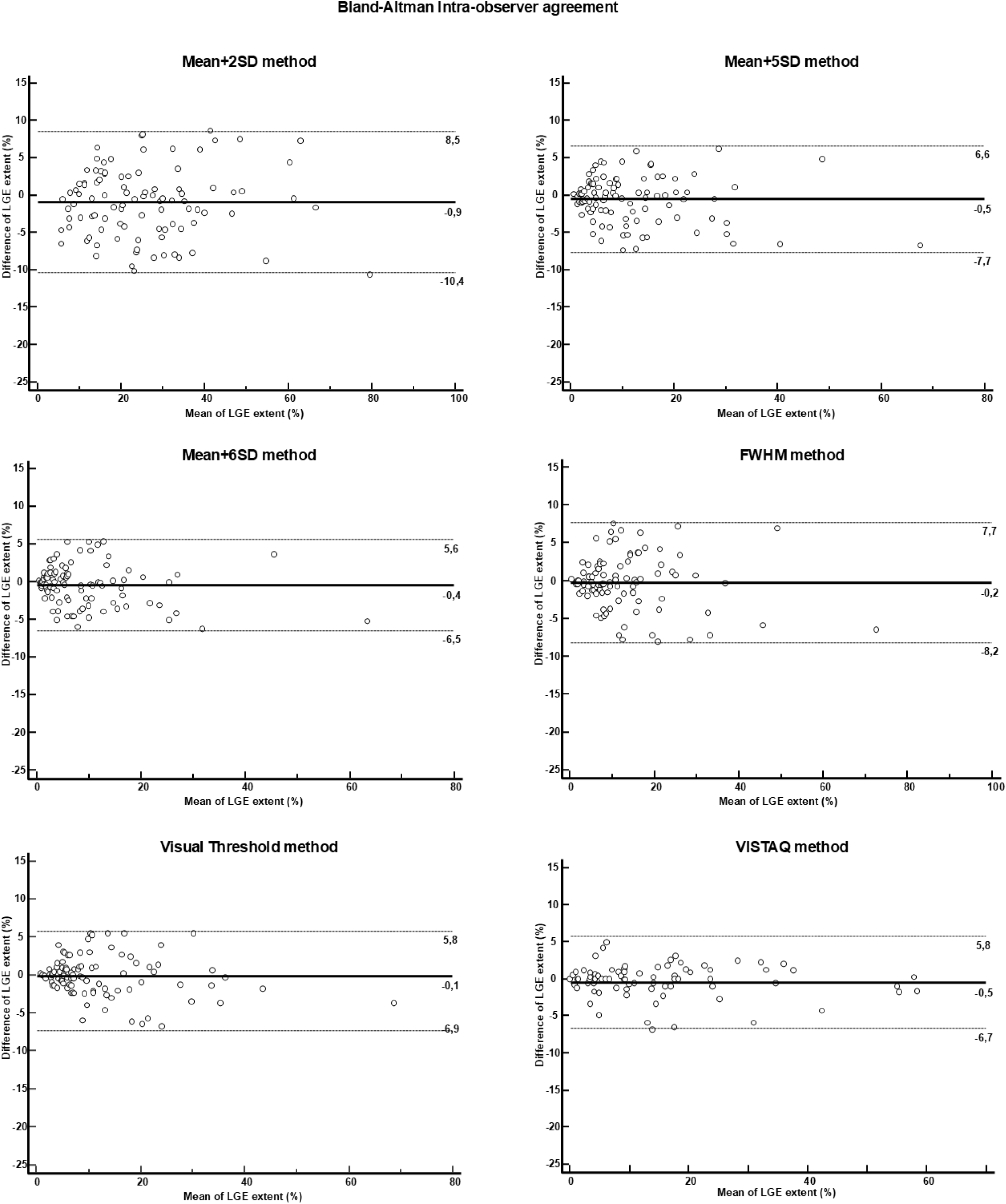
Bland-Altman plots for intra-observer agreement for quantification of LGE using different method of analysis.

**Figure 6:**
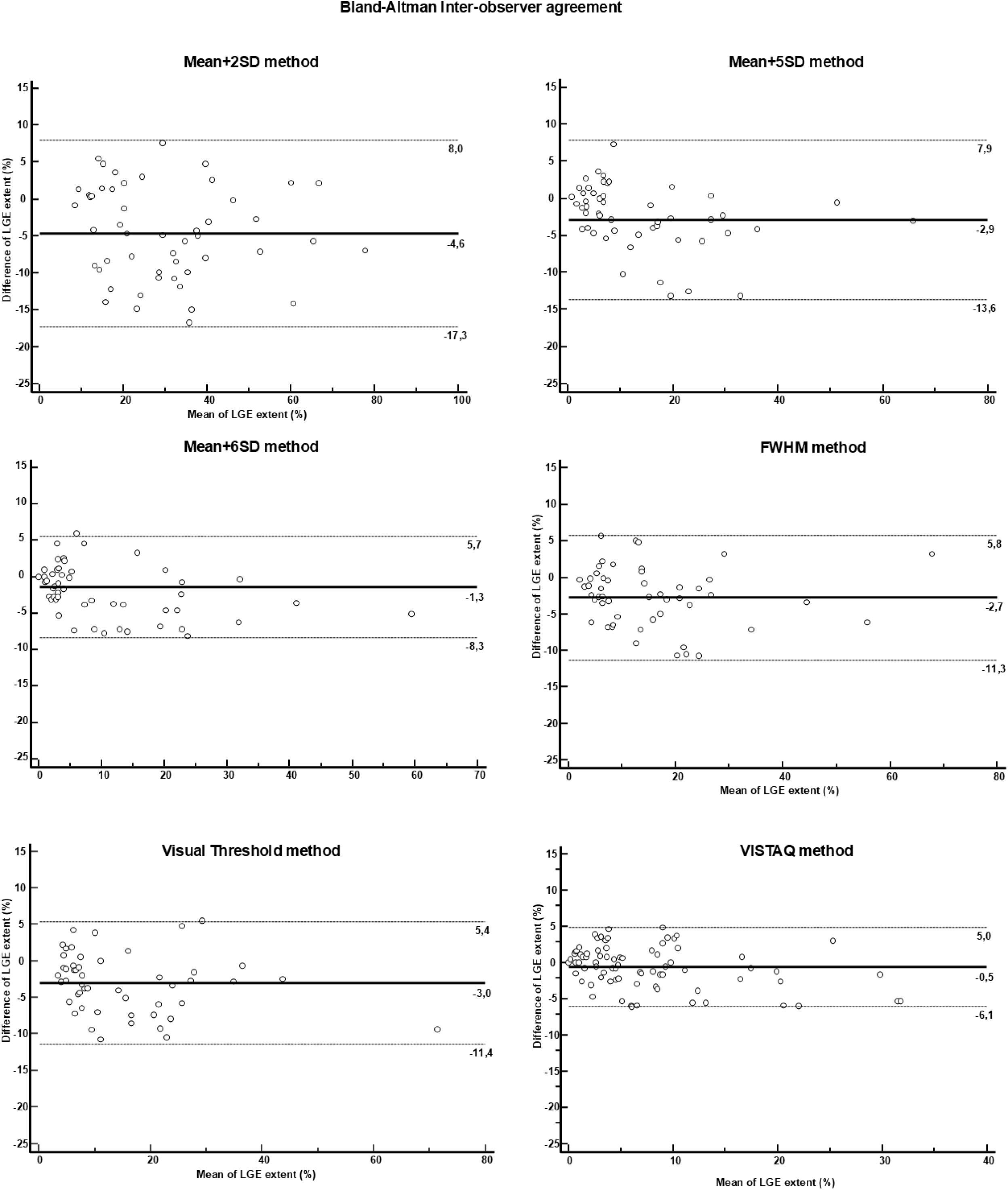
Bland-Altman plots for inter-observer agreement for quantification of LGE using different method of analysis.

### Time-duration of analysis

The median time-duration required for the analysis of LGE images with VISTAQ was significantly lower than those with the mean+2SD, +5SD+ 6SD, FWHM and manual threshold, respectively median of 105 (25-75^th^ 70-130) vs 375 (25-75^th^ 277-550) seconds, p <0.0001.

### Prognostic role of LGE extent by VISTAQ in HCM

The evaluation of prognostic role of LGE extent assessed by VISTAQ was performed in a population of 250 patients with HCM (table 5). During the median follow-up of 5 (4–10) years, hard cardiac events occurred in 21 patients (5 episodes of cardiac sudden death; 8 resuscitated cardiac arrests; 1 episode of sustained ventricular tachycardia; and 7 appropriate interventions of ICD). At ROC analysis, the best cut-off of LGE extent measured by mean+6SD for predicting hard cardiac events method was >10% with AUC of 0.75 (95% CI 0.69-0.82, sensitivity 57% and specificity 93%). The same threshold of >10% was found for LGE extent measured by VISTAQ method but with higher AUC (0.90, 95% CI 0.85-0.93), sensitivity (85%, 95% CI 64-97%) and specificity (94%, 95% 90-96%).

**Table 5.**
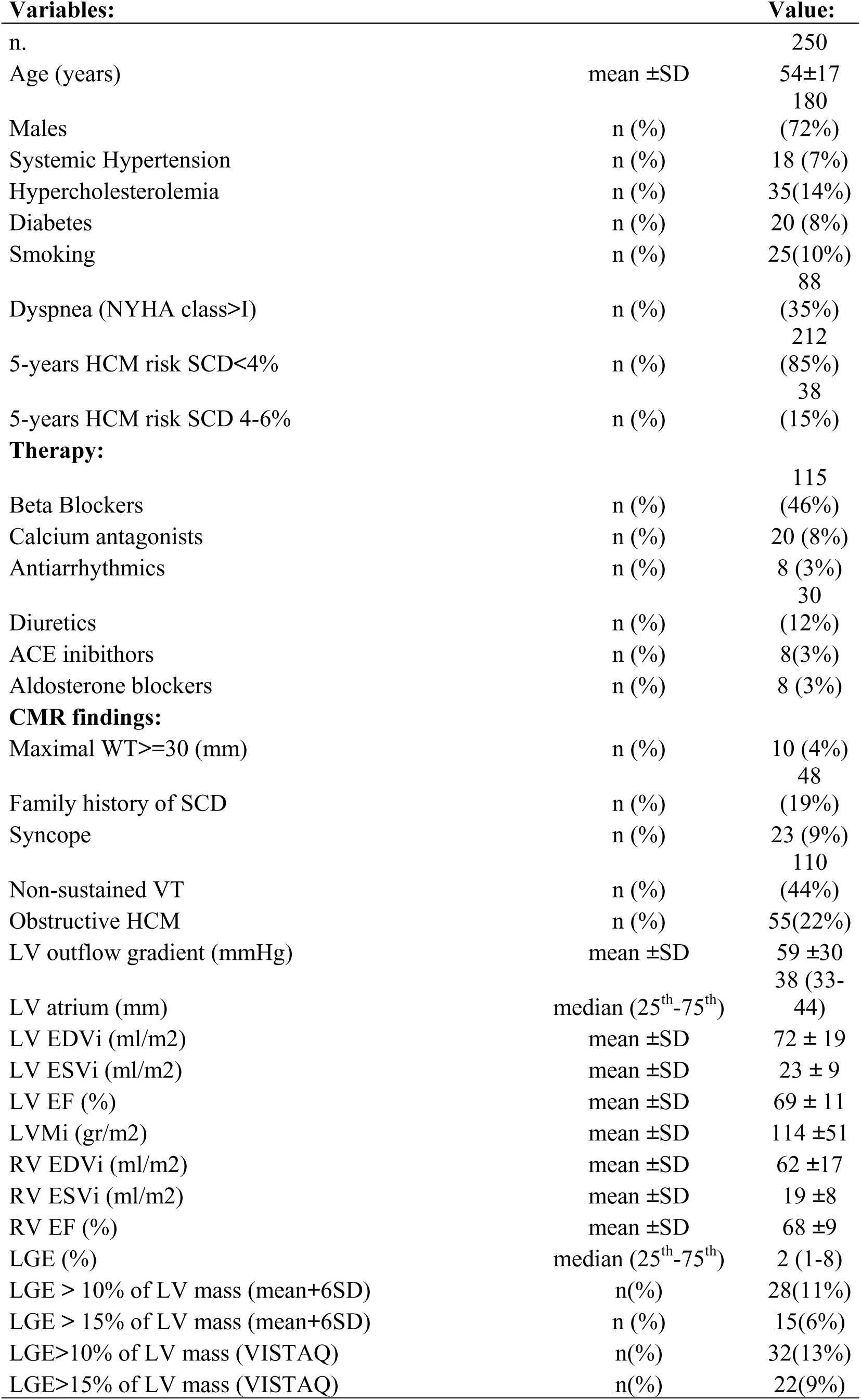
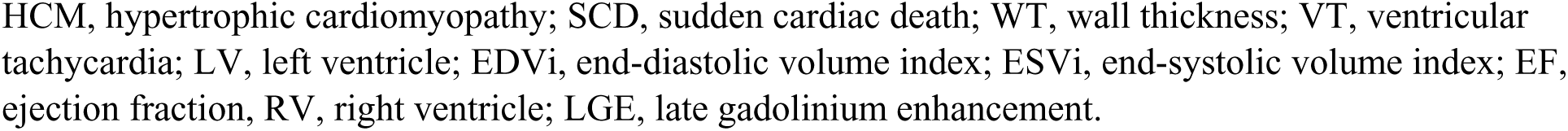
Clinical characteristics of the population of HCM for prognostic assessment of LGE extent measured by VISTAQ method.

VISTAQ and mean+6SD concordantly found a LGE extent of >10% in 23 patients, but VISTAQ found LGE extent >10% in 9 patients with lower extent by mean+6SD and on the contrary 5 patients with lower extent with VISTAQ had extent >10% using the mean+6SD technique.

The identification of patients with LGE extent >10% by VISTAQ Method allowed prediction of 18 out of 21 events, whereas when measured using the mean+6SD, patients with >10% extent had 12 events. In figure 7, Kaplan Meier survival curves for assessing survival-free from hard cardiac events are showed. Using both methods, an extent of LGE >10% was associated with worse prognosis (logrank p<0.0001 in both cases) but VISTAQ had greater HR than mean+6SD (HR 56, 95% CI 19-163, vs HR 11, 95% CI 4-32, p<0.0001).

**Figure 7:**
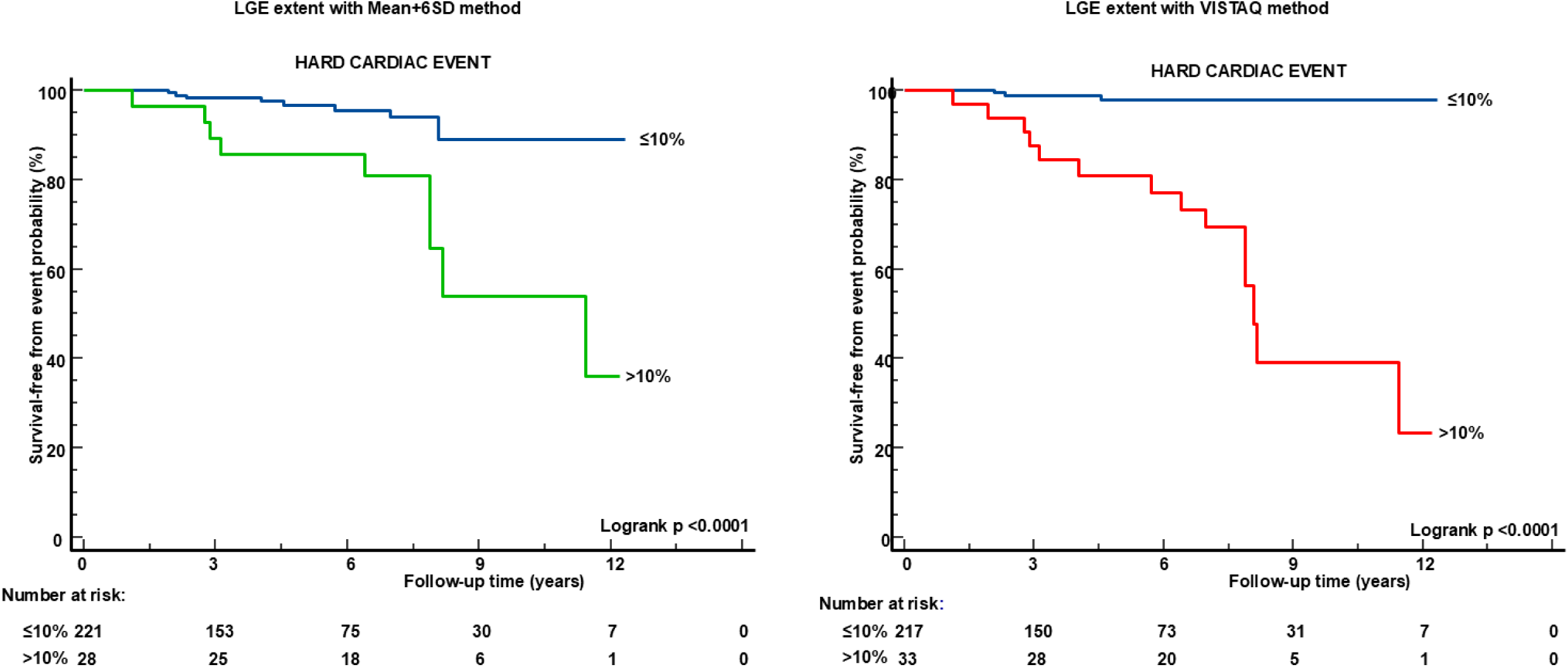
Kaplan Meier survival-free from hard cardiac events curves in patients with HCM. Patients with LGE extent >10%, measured both with mean+6SD method and both with VISTAQ, had worse prognosis than those with lower LGE extent.

## Discussion

In this study, we proposed a novel visual semiquantitative method for quantification of LGE extent, the VISTAQ which does not require myocardial contours tracing nor ROI placement.

Results may be summarized as follows: 1) VISTAQ provides excellent intra- and inter-observer reproducibility for late gadolinium enhancement (LGE) quantification across different cardiac conditions (post myocardial infarction, HCM, non-ischemic myocardial fibrosis); 2) VISTAQ had comparable reproducibility of FWHM but with lower net and absolute differences of interobserver measurement of LGE extent; 3) VISTAQ markedly shortens analysis time compared to all conventional methods; 4) quantification of LGE by VISTAQ keep its prognostic role in patients with hypertrophic cardiomyopathy (HCM).

VISTAQ is actually a semi-quantitative approach based on the analysis of myocardial mini-segments; however, with standard short-axis LGE acquisitions covering the entire left ventricle, typically yielding 100–180 mini-segments, the estimation of LGE extent closely approximates a fully quantitative assessment.

A principal strength of VISTAQ is that it eliminates the need for post-processing software, myocardial contour delineation, and placement of regions of interest in both normal and pathological myocardium. This also translates into high reproducibility. In the overall population of 250 patients, intra-observer ICC values reached 0.96 (single measures) and 0.98 (average measures), while inter-observer ICC values were 0.94 and 0.97, respectively. These values fall within the range considered excellent and indicate minimal measurement variability attributable to observer-related factors.

Importantly, reproducibility remained consistently high across distinct pathological substrates, including previous myocardial infarction, HCM, and non-ischemic LGE without left ventricular hypertrophy. This finding is clinically plausible, as LGE patterns in HCM are often patchy, intramyocardial, and less sharply demarcated compared with the subendocardial or transmural patterns typical of ischemic scar. Despite this inherent complexity, VISTAQ maintained strong consistency, underscoring its robustness even in challenging imaging phenotypes.

A possible explanation for the observed reproducibility may lie in the absence of the need to manually delineate endocardial and epicardial myocardial contours, as well as in the lack of requirement for region-of-interest (ROI) placement in either normal or pathological myocardium (as is necessary with the FWHM method).

When contouring is performed, particular care must be taken to exclude the ventricular cavity from the endocardial border. In LGE images, blood appears hyperintense due to the presence of gadolinium-based contrast agents; inadvertent inclusion of blood pool may therefore lead to overestimation of LGE extent. To avoid this, operators often trace slightly within the endocardial border. However, this approach may reduce the calculated total myocardial mass and consequently inflate the estimated LGE burden when expressed as a percentage of ventricular mass. Similarly, epicardial fat is hyperintense on LGE images, and its inclusion within the epicardial contour may also result in LGE overestimation. The need for meticulous attention to these technical aspects may increase inter- and intra-observer variability when using conventional quantification methods.

ROI placement represents a critical determinant of LGE quantification in conventional approaches, including techniques based on the mean signal intensity of remote myocardium plus 2, 5, or 6 standard deviations, as well as in the FWHM method, which requires ROI positioning within the area of maximal enhancement.

In conditions such as prior myocardial infarction, ROI placement in both remote myocardium and scar tissue is generally straightforward. In contrast, in non-ischemic cardiomyopathies, particularly hypertrophic cardiomyopathy (HCM), identification of truly unaffected myocardium may be challenging. This difficulty may also be present, although to a lesser extent, in other cardiomyopathies and myocarditis. Variability in ROI placement can lead to subtle differences in calculated mean signal intensity and standard deviation values, thereby altering the threshold definition and ultimately affecting final LGE quantification.

Furthermore, in cases of small, patchy, or highly irregular scars, such as those frequently observed in HCM or myocarditis, accurate ROI positioning within hyperenhanced regions may be difficult, representing a potential limitation of the FWHM technique.

Another relevant advantage of VISTAQ is its ability to manage imaging artifacts. With conventional quantification methods, it may be challenging to exclude false hyperintensities arising from motion artifacts, respiratory misregistration, ECG triggering issues, or suboptimal nulling of normal myocardium. These sources of error may lead to inaccurate LGE estimation when automated or semi-automated threshold-based techniques are applied.

In contrast, VISTAQ, when used by experienced operators capable of recognizing artifact-related signal abnormalities, allows these fake hyperintensities to be identified and excluded. This may potentially enable a more reliable estimation of LGE extent even in suboptimal image quality conditions.

The intra- and inter-observer reproducibility of conventional technique as mean+2SD, mean+5SD, mean+6SD, FWHM, and visual thresholding was evaluated in the present study in a cohort of patients with HCM. We chose to perform this comparison exclusively in HCM because, to date, LGE quantification is formally recommended only in this cardiomyopathy (2).

Our results are consistent with previous studies demonstrating that FWHM is the most reproducible among conventional methods (4,6). However, VISTAQ showed intra- and inter-observer ICC values comparable to FWHM, with smaller absolute and net inter-observer differences than FWHM and all other methods.

Previous studies (6–9) have evaluated only the net difference between observers, whereas in the present study we assessed both the net and the absolute differences. This distinction is methodologically relevant. When considering net differences, opposite deviations may cancel each other out. For example, if in one case the difference between observer A and B is −10% and in the subsequent case it is +10%, the mean net difference would be 0%, suggesting perfect agreement. In contrast, when absolute differences are analyzed, both values are treated as 10%, resulting in a mean absolute difference of 10%. Therefore, the assessment of absolute differences provides a more accurate estimate of the true magnitude of inter-observer variability, avoiding the potential masking effect inherent to net difference calculations.

The median net difference in LGE extent between observers was 0%, with narrow confidence intervals, while the absolute median difference was only 1.3%. All comparator techniques demonstrated significantly greater disagreement, with absolute differences ranging from approximately 2% to over 5%.

Additional noteworthy findings emerged from the comparison of LGE extent values obtained with the different methods when performed by the same operator. In the present study, consistent with previous evidence (4), the mean+2SD method proved unreliable, leading to an approximate twofold overestimation of LGE extent.

This finding is predictable and can be explained by MR physics. The distribution of signal intensity in remote (healthy) myocardium on LGE images is not Gaussian. Healthy myocardium is nulled by the inversion pulse; therefore, what is measured is not true myocardial signal but background noise. Background noise does not follow a Gaussian (normal) distribution, typical of many biological variables, but rather a Rician distribution, which is not symmetric and bell-shaped like a Gaussian curve. Instead, it exhibits a broader right-sided tail toward higher values (3). Consequently, the mean+2SD approach, which assumes Gaussian distribution, is not appropriate for LGE quantification and results in substantial overestimation.

As the number of standard deviations above the mean increases, the threshold for LGE detection shifts toward higher values, inevitably reducing the estimated LGE extent.

Our results demonstrate that the mean+6SD method, by applying a higher threshold, leads to a statistically significant, albeit modest, underestimation of LGE extent compared with mean+5SD, FWHM, visual thresholding, and VISTAQ. In contrast, measurements obtained with VISTAQ, mean+5SD, FWHM, and visual thresholding did not differ significantly from one another.

The implications of this relative “underestimation” are clinically relevant. In the multicenter study by Chan et al., in which LGE was quantified using visual thresholding, the optimal prognostic cut-off was 15% of left ventricular mass (11). Conversely, studies that quantified LGE using the 6SD method (12–14) identified an optimal cut-off of 10% of left ventricular mass. This discrepancy may, at least in part, be explained by the higher threshold and consequent underestimation of LGE extent inherent to the mean+6SD method compared with other techniques.

Lower variability of VISTAQ has important clinical implications. In risk stratification, particularly when clinical decisions may hinge on a fixed threshold of LGE extent, even small measurement differences can shift patients across risk categories. A more consistent technique therefore enhances the reliability of clinical decision-making.

Perhaps the most clinically meaningful finding of this study is the improved prognostic performance of VISTAQ in patients with HCM. Using a threshold of >10% LGE extent, VISTAQ achieved an area under the curve (AUC) of 0.90, compared with 0.75 for the mean+6SD method. This difference represents a substantial gain in discriminative ability.

Notably, sensitivity increased from 57% (mean+6SD) to 85% (VISTAQ), while specificity remained similarly high (94% vs 93%). Thus, VISTAQ identified significantly more patients who subsequently experienced hard cardiac events without increasing false positives. In absolute terms, VISTAQ correctly identified 18 of 21 events, whereas mean+6SD identified only 12.

The survival analysis further supports the clinical relevance of these findings. Although both methods showed significant prognostic separation at the >10% threshold, the hazard ratio associated with VISTAQ (HR 56) was markedly higher than that observed with mean+6SD (HR 11). While wide confidence intervals reflect the limited number of events, the magnitude of effect suggests that VISTAQ captures a biologically meaningful representation of arrhythmogenic substrate.

From a pathophysiological standpoint, myocardial fibrosis in HCM creates conduction heterogeneity and re-entry circuits, providing the substrate for malignant ventricular arrhythmias. A quantification method that more accurately and consistently captures total fibrotic burden is therefore expected to correlate more strongly with arrhythmic outcomes. The superior performance of VISTAQ may reflect a more faithful representation of this structural substrate, avoiding both overestimation from low thresholds and underestimation from excessively stringent criteria.

Beyond accuracy and reproducibility, VISTAQ demonstrated a dramatic reduction in analysis time. Median processing time was 105 seconds, compared with 375 seconds for conventional methods. This approximately 70% reduction substantially improves workflow efficiency.

Time efficiency is often overlooked in methodological studies but is critical for clinical adoption. A technique that is both highly reproducible and time-efficient is far more likely to be implemented in routine practice, particularly in high-volume imaging centers. Shorter analysis time may also reduce operator fatigue, indirectly contributing to further consistency.

### Limitations

Several limitations should be acknowledged. First, we compared VISTAQ with conventional LGE quantification methods; however, a true gold-standard reference technique is not available. As highlighted in previous studies (4), this represents an intrinsic limitation that cannot be fully overcome. Postmortem validation in animal models is inadequate due to substantial differences between human and experimental infarction patterns, the absence of reliable animal models of adult sarcomeric hypertrophic cardiomyopathy (HCM) and myocarditis, and methodological constraints inherent to autopsy studies. These include the use of non-gated (or simulated gating) imaging protocols and the impossibility of performing true post-mortem LGE imaging, which limit the comparability with in vivo clinical CMR acquisitions.

Second, although we assessed both intra- and inter-observer reproducibility, we did not evaluate inter-scan reproducibility. Additional studies specifically designed to assess scan–rescan variability are warranted to further establish the robustness of the technique.

Third, the number of hard clinical events in the prognostic analysis was relatively limited. This may have led to inflation of hazard ratio estimates and widening of confidence intervals, potentially affecting the precision of risk estimates.

Finally, although the >10% LGE threshold is consistent with prior literature, prospective studies are needed to validate the optimal prognostic cut-offs when LGE is quantified using VISTAQ.

## Conclusions

In conclusion, VISTAQ provides highly reproducible, time-efficient LGE quantification across multiple cardiac conditions and demonstrates a non-inferior prognostic performance in HCM compared with conventional threshold-based methods. If validated in larger and multicenter cohorts, VISTAQ has the potential to improve standardization of LGE assessment and refine arrhythmic risk stratification in clinical practice.

An additional advantage of VISTAQ is that this simple and rapid method, which does not require dedicated post-processing software, may be directly applied by clinical cardiologists to independently reassess LGE extent. This could allow clinicians to personally verify and, if necessary, recalculate LGE burden in order to optimize patient management, rather than relying exclusively on the values reported in the formal CMR report.

## Data Availability

The data that support the findings of this study are available from the corresponding author upon reasonable request.

